# COVID-19 Cases and Testing in 53 Prison Systems

**DOI:** 10.1101/2020.08.25.20181842

**Authors:** Katherine Lemasters, Erin McCauley, Kathryn Nowotny, Lauren Brinkley-Rubinstein

**Author notes:** Corresponding Author: Lauren Brinkley-Rubinstein, PhD, 333 S. Columbia Street, Chapel Hill, NC 27559, Lauren.

## Abstract

**Background:** COVID-19 has entered United States prison systems at alarming rates. Disparities in social and structural determinants of health disproportionately affect those experiencing incarceration, making them more vulnerable to COVID-19. Additionally, prisons are sites of congregate living, making it impossible to practice social distancing, and most prisons have relied only on incremental measures to reduce risk and spread of COVID-19. To more fully understand the impact that COVID-19 is having on incarcerated populations, it is critical to have systematic data on testing, test positivity, cases, and case fatality. Using data from the COVID Prison Project, we present data on 53 prison systems COVID-19 testing, test positivity, case, and case fatality by state and compare these data with each state’s general population.

**Results:** Many states are not reporting full information on COVID testing with some also not reporting on case fatality. Among those reporting data, there is a wide variation between testing, test positivity, and case rates within prison systems and as compared to the general population. However, when more tests are deployed more cases are identified with the majority of state prisons having higher case rates than their general population.

**Conclusions:** These findings underscore the need for the implementation and study of COVID-19 mitigation and surveillance strategies to flatten the COVID-19 curve in prisons across the country. We call for future research to build on this data from the COVID Prison Project to protect the health of our nations’ often forgotten residents.

## Introduction

The United States (US) leads the world in both COVID-19 and mass incarceration. In the US, to date, there are 3,475,698 infections and 136,649 deaths in the US.^1^ While the US has only 4% of the world’s population, it has a quarter of the world’s COVID-19 infections and deaths.^1,2^ Simultaneously, the US has over a quarter of the world’s incarcerated population.^3^ Yet, these two public health emergencies do not exist in silos. COVID-19 has entered our country’s prisons, jails, and communities most impacted by mass incarceration at astounding rates.^4,5^ In fact, jails and prisons have become epicenters of the pandemic with nine of the ten largest single site outbreaks occurring in correctional settings.

The confluence of mass incarceration and COVID-19 is not a coincidence. Disparities in social and structural determinants of health disproportionately impact racial minorities, those experiencing homelessness, and persons with substance use disorders or mental illness, all of whom are more likely to be incarcerated due to mass incarceration’s roots in racial inequities and discriminatory practices.^6,7^ People who are incarcerated also have a higher burden of poor health outcomes making them more likely to suffer severely from COVID-19.^7^ Carceral settings themselves also amplify the risk of COVID with common prevention activities like social distancing being impossible to engage in due to dormitory style living conditions or overcrowding.^7^ In addition, the communities that people return home to and come into prisons from also bear a disproportionate burden of COVID-19. Black Americans are both overrepresented in the CJ system and are more likely to live in areas with higher poverty rates, have limited health care access, and have higher rates of jobs in service industries where they are less able to work from home, which increases exposure to COVID-19.^8,9^

Policymakers, advocates, and academics have called for decarceration, specifically by increasing the number of compassionate releases, eliminating cash bail, and releasing those imprisoned for low-level crimes and misdemeanors.^7,10^ However, prisons have yet to do this, instead relying on incremental measures such as suspending family and legal visits, distributing masks, and reducing the amount of time that people can spend recreating.^11^ On average, prisons have only reduced their populations by 5%.^12^ In the meantime, outbreaks have continued to occur in prisons throughout the country.

Given these outbreaks and the vulnerability of the imprisoned population, it is important that we comprehensively document the prevalence of COVID-19 within prisons. To do this, it is critical that we have systematic data on testing, test positivity, cases, and case fatality. While this is a pressing national issue, this data must be assessed at the state level as each state’s Department of Corrections collects data independently and independently determines their COVID policies. Additionally, each state’s general population has a different prevalence of COVID-19 infection and differing COVID policy (e.g., mandatory masks; stay-at-home orders), making the context surrounding this data unique to each state. In this paper, we present testing, infection, and fatality data by state, using novel data from the COVID Prison Project (CPP) on each state prison’s COVID data. We call for future research to build on CPP data to protect the health of our nations’ often forgotten residents.

## Methods

### COVID-19 Prison Data

For COVD-19 data in prisons, we used publicly accessible data from the (CPP).^4^ The CPP publishes a daily aggregate dataset examining COVID-19 in correctional facilities, including data on the number of tests, the number of confirmed positive cases, and mortality, among other factors, due to COVID-19 among correctional staff and incarcerated individuals. The data are aggregated by CPP based on public reports by prison systems. Each day, counts were extracted from Department of Corrections websites and supplemented with information from media reports and press releases. In total the CPP collects daily data for 53 prison systems (all 50 states, Puerto Rico, the Federal Bureau of Prisons, and Immigration and Customs Enforcement (ICE).

### Prison Population Data

For prison population data, we used data collected by the Vera Institute of Justice.^13^ Prison population counts were collected during the first quarter of 2020, and largely capture the changes in population size due to COVID-19. When possible, we use the most recent count (collected on April 30th / May 1st, 2020). For states without data from this most recent wave, we use the second most recent count (December 31st for Illinois, Maryland, Minnesota, New Mexico and Virginia; and March 31st for Montana, South Dakota, Tennessee, and Washington). For ICE, we use the currently detained population as of July 11^th^ 2020.^14^ For Puerto Rico, we use data from the Bureau of Justice Statistics, last updated in 2012.^15^

### General Population Data

For general population COVID-19 counts, we used data for positive cases and fatalities from the Johns Hopkins Coronavirus Resource Center and data on testing numbers from the COVID Tracking Project.^1,2^ For the general population count, we used data from The U.S. Census Bureau.^16^ The U.S. Census Bureau collects annual population estimates by state in July of each year. This study uses the 2019 state population estimates to create rates for the general population. Data come from all 50 US states and Puerto Rico. COVID-19 cases were captured cumulatively and include active cases, recovered cases, and fatalities. COVID-19 tests and fatalities were also captured cumulatively. General population COVID-19 data on case numbers and fatalities are available through both the John Hopkins Coronavirus Resource Center and the COVID Tracking Project. As a sensitivity analysis we calculated rates using both sources and found similar patterns. Alternative tables using the COVID Tracking Project data on cases and fatalities are available by request.

### Analysis

We calculated crude COVID-19 testing, case, and case fatality rates separately by state for both the prison and general population. We also calculated the test positivity (the percent of tests that are returned positive) in the prison population and general population by state. Data is presented at the state level due to large differences in both testing strategies and COVID-related policies in each state. The World Health Organization recommends test positivity being between 3-12% based on positivity rates from countries that have successfully mounted extensive testing programs.^17^ This is the standard that we use.

## Results

As of July 15, 2020, 10 states and Puerto Rico were reporting no testing information (Table 1). Of those that report testing numbers, testing varies widely with Hawaii testing 6 per 1,000 people who are incarcerated and Minnesota testing 1,531 per 1,000 people who are incarcerated, indicating an average of more than one test per incarcerated person. Maryland, Massachusetts, Michigan, Minnesota, Missouri, New Jersey, Rhode Island, Tennessee, Texas, Vermont, West Virginia, and Wisconsin have all administered more than 500 tests per 1,000 incarcerated people, with Maryland, Massachusetts, Michigan, Minnesota, and Vermont having all administered more than one test per incarcerated person-indicating a universal testing effort in which all people currently incarcerated have had access to a COVID-19 test. The majority of prison systems have tested a higher proportion of their incarcerated population than the state’s general population. However, eleven prison systems are still testing fewer incarcerated people per 1,000 than their state’s general population.

**Table 1.**

| State | Tests per 1,000 |  | Positive Cases per 1,000 |  | Fatalities per 1,000 Cases |  | Test Positivity Rate (%) |  |
| --- | --- | --- | --- | --- | --- | --- | --- | --- |
|  | General Population | Prison Population | General Population | Prison Population | General Population | Prison Population | General Population | Prison Population |
| Alabama | 110 | 21 | 12 | 5 | 21 | 9 | 11 | 24 |
| Alaska | 213 | 400 | 2 | 2 | 10 |  | 1 | 0 |
| Arizona | 101 | 109 | 18 | 14 | 19 | 2 | 18 | 13 |
| Arkansas | 131 | 406 | 10 | 149 | 11 |  | 8 | 37 |
| California | 147 | 443 | 9 | 55 | 21 | 1 | 6 | 12 |
| Colorado | 71 | 357 | 7 | 36 | 46 | 0 | 9 | 10 |
| Connecticut | 169 |  | 13 | 122 | 92 | 1 | 8 |  |
| Delaware | 147 |  | 13 | 55 | 40 | 3 | 9 |  |
| Florida | 127 | 308 | 14 | 28 | 15 | 1 | 11 | 9 |
| Georgia | 109 |  | 12 | 15 | 24 | 3 | 11 |  |
| Hawaii | 69 | 6 | 1 | 0 | 17 | 0 | 1 | 0 |
| Idaho | 75 | 180 | 7 | 76 | 9 |  | 9 | 42 |
| Illinois | 164 | 24 | 12 | 9 | 47 | 4 | 8 | 37 |
| Indiana | 87 | 79 | 8 | 27 | 52 | 3 | 9 | 35 |
| Iowa | 123 | 450 | 12 | 25 | 21 | 0 | 9 | 6 |
| Kansas | 82 |  | 7 | 93 | 15 | 0 | 9 |  |
| Kentucky | 103 |  | 5 | 29 | 31 | 1 | 5 |  |
| Louisiana | 215 | 55 | 18 | 22 | 41 | 2 | 8 | 41 |
| Maine | 95 | 588 | 3 | 2 | 32 |  | 3 | 0 |
| Maryland | 112 | 1024 | 12 | 34 | 45 | 1 | 11 | 3 |
| Massachusetts | 143 | 1010 | 16 | 54 | 74 | 2 | 11 | 5 |
| Michigan | 137 | 1017 | 8 | 103 | 80 | 2 | 6 | 10 |
| Minnesota | 140 | 1531 | 8 | 39 | 36 | 1 | 6 | 3 |
| Mississippi | 120 | 15 | 13 | 5 | 33 | 1 | 11 | 31 |
| Missouri | 84 | 618 | 5 | 10 | 37 | 0 | 6 | 2 |
| Montana | 116 | 404 | 2 | 0 | 16 |  | 2 | 0 |
| Nebraska | 114 | 162 | 11 | 1 | 13 |  | 10 | 1 |
| Nevada | 118 |  | 10 | 1 | 20 |  | 8 |  |
| New Hampshire | 101 | 17 | 4 | 0 | 64 |  | 4 | 2 |
| New Jersey | 196 | 714 | 20 | 159 | 89 | 2 | 10 | 22 |
| New Mexico | 205 | 218 | 8 | 24 | 35 | 1 | 4 | 11 |
| New York | 249 | 99 | 21 | 13 | 80 | 3 | 8 | 13 |
| North Carolina | 122 | 268 | 9 | 32 | 17 | 1 | 7 | 12 |
| North Dakota | 166 |  | 6 | 5 | 19 | 0 | 4 |  |
| Ohio | 91 | 346 | 6 | 105 | 44 | 2 | 7 | 30 |
| Oklahoma | 113 | 164 | 6 | 0 | 19 |  | 5 | 0 |
| Oregon | 72 | 112 | 3 | 22 | 19 | 0 | 4 | 20 |
| Pennsylvania | 76 | 154 | 8 | 6 | 68 | 4 | 10 | 4 |
| Puerto Rico** | 64 |  | 3 | 0 | 16 |  | 5 |  |
| Rhode Island | 166 | 701 | 17 | 10 | 56 | 0 | 10 | 1 |
| South Carolina | 102 |  | 12 | 31 | 16 | 1 | 12 |  |
| South Dakota | 106 | 33 | 9 | 1 | 15 | 0 | 8 | 3 |
| Tennessee | 161 | 894 | 10 | 122 | 11 | 0 | 6 | 14 |
| Texas | 91 | 912 | 10 | 80 | 12 | 1 | 11 | 9 |
| Utah | 135 | 87 | 10 | 1 | 8 | 0 | 7 | 1 |
| Vermont | 126 | 1031 | 2 | 34 | 42 |  | 2 | 3 |
| Virginia | 99 |  | 9 | 42 | 27 | 1 | 9 |  |
| Washington | 94 | 156 | 6 | 15 | 33 | 1 | 6 | 9 |
| West Virginia | 122 | 771 | 3 | 19 | 22 |  | 2 | 2 |
| Wisconsin | 125 | 1068 | 7 | 13 | 21 |  | 6 | 1 |
| Wyoming | 74 |  | 3 | 0 | 11 |  | 5 |  |
| BOP |  | 172 |  | 52 |  | 11 |  | 30 |
| ICE* |  | 607 |  | 157 |  | 1 |  | 26 |

### Testing Positivity

Among those tested, test positivity varied by state from 0-42%. Systems with the highest test positivity have, on average, tested a small proportion of their incarcerated population. For example, Louisiana has tested 55 per 1,000 incarcerated people and has a test positivity of 41%. Similarly, Idaho has tested 180 per 1,000 incarcerated people and has a test positivity of 42%. Test positivity is, on average, higher in prison systems than the general population, but some prison systems have lower test positivity than their general population. This includes states with recent outbreaks in their general population (e.g., Arizona, Florida), those testing very few people who are incarcerated (e.g., Nebraska), and those conducting mass testing (e.g., Minnesota).

### Case Rates and Case Fatality Rates in Prisons and the General Population

Thirty-four of the prison systems have case rates per 1,000 that are higher than the general population. New Jersey has the highest cases per 1,000 incarcerated people at 159/1,000. Among those detecting more than 100 cases per 1,000 incarcerated people (Arkansas, Michigan, New Jersey, Ohio, Tennessee), all have tested a high proportion of their population. By comparison, the highest case rate in the general population is in New York with 21 per 1,000 testing positive. Six states (Arkansas, Connecticut, Michigan, New Jersey, Ohio, and Tennessee) have detected over 100 cases per 1,000 greater than their state’s general population. For instance, New Jersey has detected 159 cases per 1,000 incarcerated people and 20 cases per 1,000 in the general population, resulting in a disparity of 139 cases per 1,000 individuals in the state of New Jersey among those in prison. Thirty-seven states are reporting case fatality data. Fatalities per 1,000 cases range from zero in Hawaii, North Dakota, Rhode Island, South Dakota, and Utah to 11 deaths per 1,000 cases in the Federal Bureau of Prisons.

## Discussion

This analysis indicates that there is wide variation between testing and case rates in various prison systems and the general population. Previous research has estimated national prevalence of various state and federal prison systems.^18^ However, it is important to understand the COVID-19 pandemic and its impact on prison systems at a more granular level. For instance, when pooling low incidence prison settings with those in the middle of an outbreak disguises the local effect that COVID-19 is having on states and communities across the country. Our findings demonstrate that more often than not, more testing equals more cases identified, but uneven testing rates across the states make opaque a clear understanding of the extent of COVID-19 incidence in prison settings in several states.

Decarceration must remain a priority as it is the most effective way to prevent COVID-19 in carceral settings. In concert, though, prison systems should be the focus of robust testing efforts and be provided the resources to develop and deploy long-term testing strategies. The Centers for Disease Control and Prevention has indicated that asymptomatic testing in prison settings is warranted.^19^ A number of different testing strategies that should be implemented and studied relevant to their efficacy. These include: targeted testing (e.g. testing all new entrants, all transfers); public health surveillance testing (e.g. sampling a proportion of each facility or housing unit who are at high risk and testing them every month); and universal testing (repeat testing of all incarcerated people in predefined intervals over time). Not to be overlooked is the importance of implementing staff testing strategies as well.

Underlying this conversation is the need for updated, public information on COVID spread within our country’s prisons. This includes integrating data on COVID-related policies in prisons and their surrounding communities, keeping Departments of Corrections accountable by mapping their testing strategies to actual testing, and measuring the impact of decarceration, where it does occur, on COVID spread. As the pandemic of COVID persists, it is increasingly crucial that we collect and analyze this data to ultimately flatten the COVID-19 curve in prisons.

### Limitations

One limitation of this analysis is that the prison population is demographically distinct from that of the general population. We do not present standardized estimates because this information is not available at the state level (i.e., COVID-19 deaths by age, sex, and race) and the data which is available is likely systematically biased due to COVID-19 releases which prioritize high risk populations. It is important to keep in mind that given mass incarceration’s racist roots, prison populations have a higher percentage of Black inmates than there are Black Americans in the general population.^6^ While 13% of the US population is Black, 33% of inmates are.^20,21^ Prisons also have a much smaller percentage of females (7%) than in the general population (51%), and a smaller percentage over the age of 65 (3%) than in the general population (17%).^21,22^ Additionally, the data available for this analysis is publicly reported by prison systems. Accuracy of reporting and real-time reporting were outside the control of the research team.

## Conclusion

The two pandemics of mass incarceration and COVID are increasingly interconnected. COVID-19 cases are highly concentrated in prison systems. This study found that testing and case rates vary widely from state to state, but, overall, more testing leads to more case identification. To accurately assess COVID-19’s impact in the US prisons, it is urgently important to implement long-term testing strategies. It is only through implementing these strategies and continually documenting COVID testing, cases, and fatalities that we will know the spread of COVID in our prison and the injustices it has created and perpetuated.

## Data Availability

The datasets generated and/or analyzed during the current study related to prison COVID data are available in the COVID Prison Project repository, https://covidprisonproject.com. Datasets relate generated and/or analyzed during the current study related to the prison's population are available at the Vera Institute of Justice. Datasets relate generated and/or analyzed during the current study related to the general population and COVID are available at the Johns Hopkins Coronavirus Resource Center. Datasets relate generated and/or analyzed during the current study related to the general population are available at the US Census Bureau.

